# Polygenic risk scores for nicotine use and family history of smoking are associated with smoking behaviour

**DOI:** 10.1101/2022.12.19.22283408

**Authors:** Jerome C. Foo, Fabian Streit, Josef Frank, Norman Zacharias, Lea Zillich, Lea Sirignano, Maja P. Völker, Peter Nürnberg, Thomas Wienker, Michael Wagner, Markus Nöthen, Michael Nothnagel, Henrik Walter, Bernd Lenz, Rainer Spanagel, Falk Kiefer, Georg Winterer, Marcella Rietschel, Stephanie H. Witt

**Author notes:** **Correspondence To:** Stephanie H. Witt, PhD, Department of Genetic Epidemiology in Psychiatry, Central Institute of Mental Health, Medical Faculty Mannheim, University of Heidelberg, Mannheim, Germany.

## Abstract

**Introduction:** Formal genetics studies show that smoking is influenced by genetic factors; exploring this on the molecular level can offer deeper insight into the etiology of smoking behaviours.

**Methods:** Summary statistics from the GWAS and Sequencing Consortium of Alcohol and Nicotine (GSCAN) Consortium were used to calculate polygenic risk scores (PRS) in a sample of ∼2,200 smokers/never-smokers. The association of PRS for Smoking Initiation (i.e. Lifetime Smoking; SI-PRS) with smoking status, and PRS for Cigarettes per Day (CpD-PRS) and Smoking Cessation (SC-PRS) with Fagerström Test for Nicotine Dependence (FTND) score were examined, as were distinct/additive effects of parental smoking on smoking status.

**Results:** SI-PRS explained 6.65% of variance (Nagelkerke-R^2^) in smoking status (p=1.71×10^−24^). In smokers, CpD-PRS (R^2^=3.15%, p=1.82×10^−8^) and SC-PRS (R^2^=2.01%; p=7.18×10^−6^) were associated with FTND score. Parental smoking alone explained R^2^=3.06% (p=2.43×10^−12^) of smoking status, and 1.39% when added to the most informative SI-PRS model (total R²=8.04%).

**Conclusion:** These results show the potential utility of molecular genetic data for research investigating smoking prevention. The fact that PRS explains more variance than family history highlights progress from formal to molecular genetics; the overlap and increased predictive value when using both suggests the importance of combining these approaches.

**Implications:** This study underlines the value of using PRS to predict smoking status/behaviour. It highlights the importance of molecular genetic methods in research investigating smoking prevention and points to the necessity of combining family history and molecular genetic data.

## Introduction

Smoking behaviours are related to a multitude of diseases including cancer, chronic obstructive pulmonary disease (COPD), and heart disease. Despite the fact that its adverse effects on health are well-known and considerable efforts to lower its prevalence have been made by many countries, the global number of smokers is still rising ^1^, leading to increasing preventable mortality worldwide.

Twin and family studies have shown that smoking behaviour is influenced by genetic factors ^2-4^, and it is thought that a large proportion of variance in smoking-related behaviours is accounted for by genetic effects ^5^. The different stages of smoking (initiation, regular smoking, nicotine dependence, cessation) have strong genetic components which overlap ^5,6^. Better understanding of the genetic factors involved in the development of smoking is needed to take more effective steps towards prevention.

One method of exploring the influence of genetic load on phenotypes is polygenic risk score (PRS) profiling, which takes genetic variants identified in GWAS as being associated with a given phenotype and combines them into a polygenic risk score that captures part of an individual’s susceptibility to developing that phenotype ^7,8^. In polygenic risk scoring, summary results of risk variants and effect sizes identified in GWAS of particular phenotypes are used as so-called “training samples”, and used to generate risk scores reflecting the genetic risk burden for the phenotype in an independent “target sample”. PRS are beginning to show utility not only in research-based case control studies, but also at the level of population-based cohort studies ^7^. While it has been suggested that PRS are beginning to have clinical utility in terms of disease risk prediction ^7^, for many phenotypes, predictive ability remains modest, and more evidence is still needed on the value of adding PRS to prediction scenarios in clinical settings ^9^.

Family history is another measure capturing genetic risk, but also comprises environmental familial influences ^10^. Several recent studies have started to explore the interplay of genome-wide PRS and family history, finding that for a variety of common diseases, the two measures are complementary, partially independent and not interchangeable ^11^.

In the present study, we aimed to explore the influence of genetic load on smoking phenotypes by investigating whether PRS of nicotine phenotypes could predict smoking status and intensity of dependence, and how this compared to prediction using family history. PRS were calculated based on summary statistics from the largest GWAS of nicotine phenotypes (cigarettes per day, smoking initiation) to date^6^, which did not include the present sample, and were used to predict smoking status and behaviour in a population-based case-control study of smokers and never-smokers (n= 2,396) ^12^.

## Methods

### Data Collection

Demographic and genetic data from the population-based German multi-centre study “Genetics of Nicotine Dependence and Neurobiological Phenotypes” (German Research Foundation, SPP1226) were used. Full details on the data collection in the cohort can be found in Lindenberg et al. 2011^12^. Briefly, in 2007-2009, smokers (n=1,116) and never-smokers (n=1,280) aged 18-65 years were recruited from the general population. Exclusion criteria included being a former smoker, alcohol or substance abuse/dependence, non-German ethnicity, pregnancy, and medical conditions or medication that might interfere with the study.

Classification of smokers was conducted using DSM-IV criteria, SCID and the German version of the Fagerström Test for Nicotine Dependence (FTND)^12^. FTND scores were available for 2,228 individuals (n=1,027 smokers, n=1,201 never-smokers). Parental history of smoking was assessed via questionnaire.

### Genotyping and Quality control of the Target Sample

Genetic data was available from 2,312 individuals. DNA was extracted from whole blood and prepared using the Qiagen FlexiGene DNA Kit (Qiagen, Hilden, Germany) according to manufacturer protocols. Genotyping was performed using the Infinium OmniExpress Exome Array (Illumina).

Quality control was performed using PLINK 1.90 ^13^. Individuals were excluded for being ancestry outliers (>4.5 standard deviations on any of the first 20 ancestry principal components), relatedness (Pi-HAT>0.2) and person–missing rate (≥ 0.02); Single nucleotide polymorphisms (SNPs) were excluded based on missing rate (≥ 0.02), deviation from Hardy-Weinberg disequilibrium (HWE–P-value ≤ 1×10^−6^), and minor allele frequency (< 0.01).

The final data set available for PRS analyses contained n=2,169 individuals (997 smokers, 1172 never-smokers) and 652,021 SNPs.

### Polygenic Risk Scoring and Summary Statistics

GWAS meta-analysis summary statistics from the GWAS and Sequencing Consortium of Alcohol and Nicotine (GSCAN) excluding the 23andme subset were used, comprising the phenotypes: 1) smoking initiation (i.e. ever being a smoker vs. never; SI; n=632,802); 2) cigarettes per day (CpD; n=263,954); and 3) smoking cessation (SC; n=312,821). Details on the phenotype definitions in GSCAN can be found in Liu et al. 2019 ^6^ and at https://genome.psych.umn.edu/images/d/da/GSCAN_GWAS_Phenotype_Definitions-2-24-2016.pdf (Accessed May 5, 2022).

PRS were calculated in PRSice 2 ^14^, and regression analyses were performed in R 3.6.3. The extended MHC region was excluded from calculations. Clumping was carried out using a threshold of pairwise r^2^= 0.1 with a maximum distance of 250kb. To optimise prediction, PRS were calculated for a range of p-value thresholds, e.g., for each keeping only SNPs with a GWAS association P-value below the threshold (P_t_: 5×10^−8^, 1×10^−6^, 1×10^−4^, 0.001, 0.01, 0.05, 0.1, 0.2, 0.5, 1.0).

The smoker/never-smoker dataset described above was used as the target sample. Regression analyses were used to test for association of PRS of phenotypes of interest (at all thresholds above) with smoking status (smokers vs. never-smokers: SI), and FTND score (in smokers; CPD, SC).

Analyses were adjusted for population stratification by including the first 10 ancestry principle components. R^2^ (Nagelkerke pseudo R² for categorical phenotypes) was used to estimate explained variability. Model fits were calculated as R^2^ of the full model (e.g. Pheno ∼ PRS + PCs) and -R^2^ of the null model (Pheno ∼ PCs).

In an additional step, specifying an incremental variable in the regression, we investigated the contribution of family history of smoking (for SI-PRS: parental history of smoking, 0=neither, 1=one, 2=both parents).

## Results

### Smoking status

PRS for SI explained R^2^=6.65% of variance in smoking status (at the most informative threshold of Pt=0.5, p=1.71×10^−24^) (yellow-orange-red, **Fig.1a**). Quantile plot shows the increasing effect of higher SI-PRS (**Fig. 2a**).

**Figure 1.**
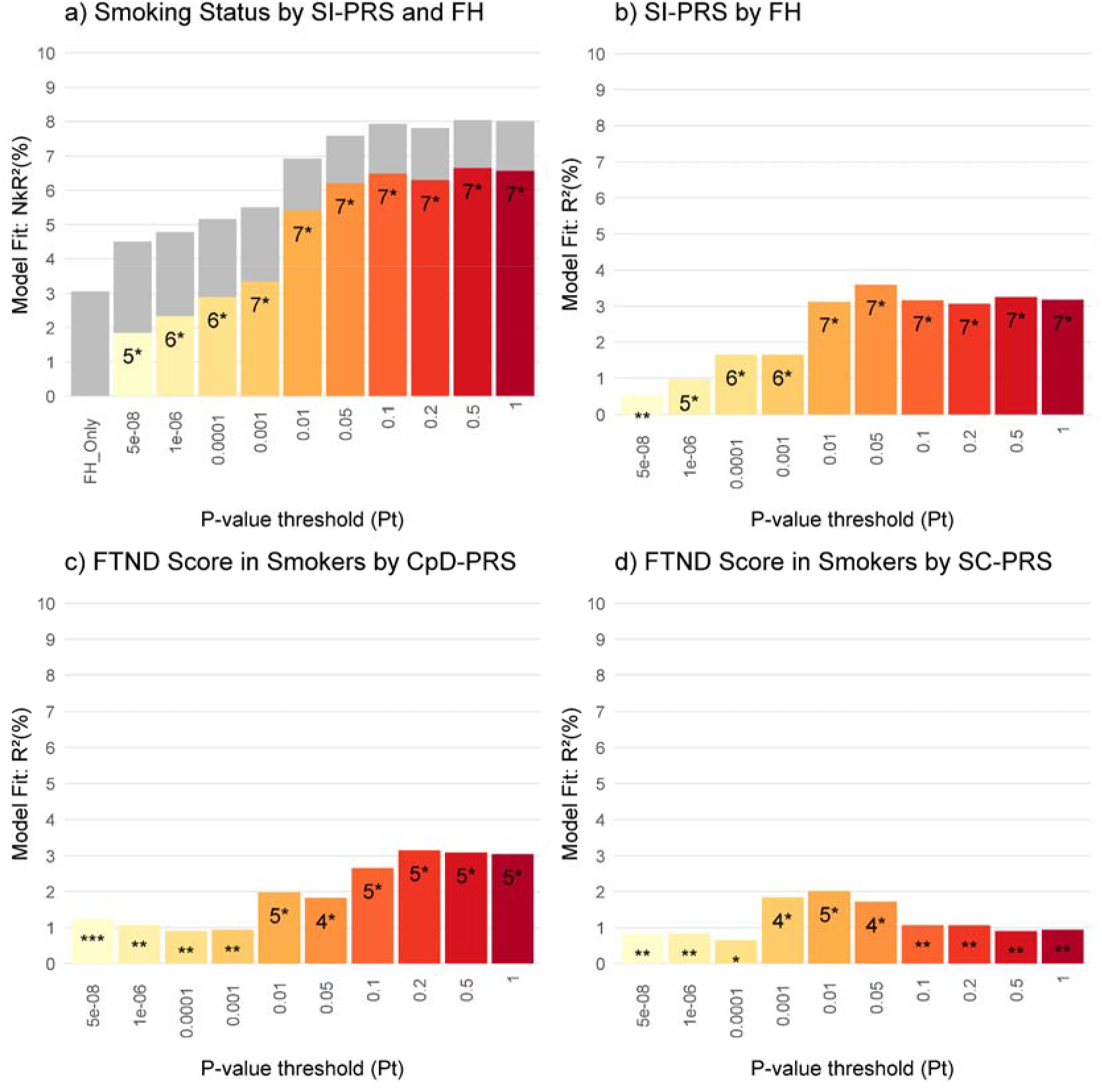
**a)** Smoker/Never-smoker Status Predicted by Parental Smoking (gray) and PRS for SI (colored bars); **b)** Association of Parental Smoking with PRS for SI at all p-value thresholds**; c)** FTND Score Predicted by PRS for CpD**; d)** FTND Score Predicted by PRS for SC. Abbreviations *PRS: Polygenic Risk Scores; FH: Family History of Parental Smoking; FTND: Fagerström Test for Nicotine Dependence; SI: Smoking Initiation; CpD: Cigarettes per Day; SC: Smoking Cessation; NkR*^*2*^: *Nagelkerke’s R*^*2*^. *\*p<0*.*05, **p<0*.*005, ***p<0*.*001, 4*p<0*.*0001, 5*p<0*.*00001, 6*p<0*.*00000001, 7*p<0*.*000000000001*.

**Figure 2.**
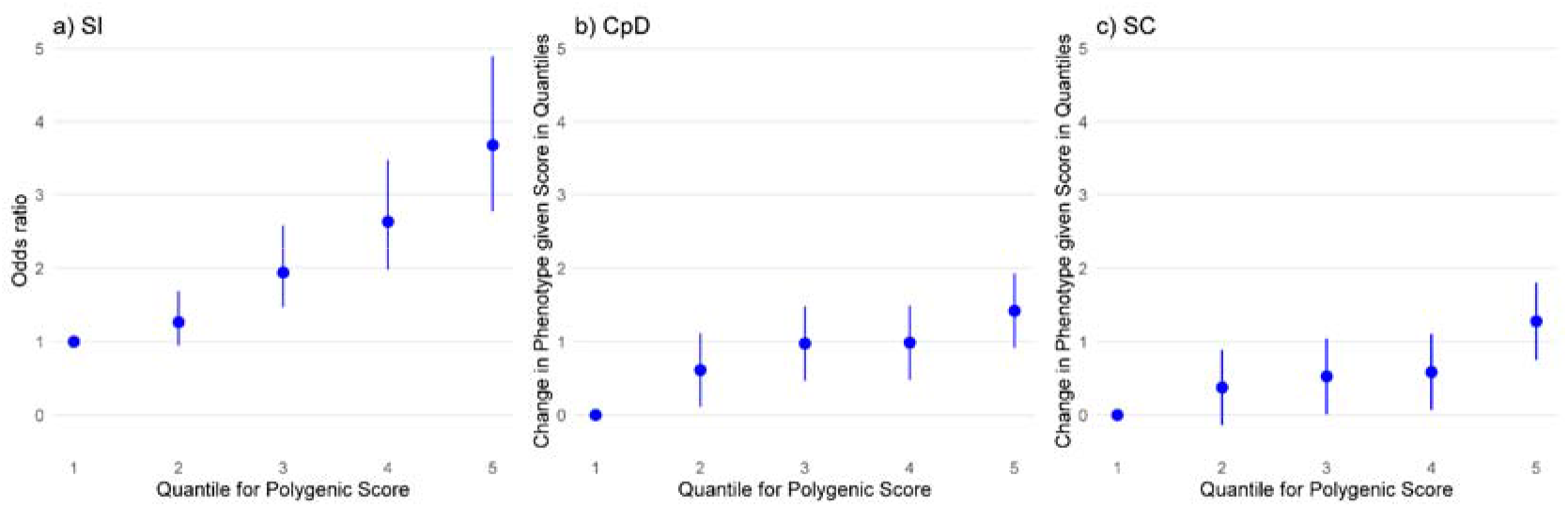
Quantile plots showing: **a)** Odds Ratio for SI-PRS on smoker/non-smoker status; **b)** Change in FTND score given CpD-PRS and **c)** Change in FTND score given SC-PRS. 1 is the reference quintile in all plots..

Parental Smoking alone explained R^2^=3.06% (p=2.43×10^−12^) of smoking status. In the model combining parental smoking and PRS, at the most informative PRS threshold, parental smoking explained an additional and independent R^2^=1.39% of variance (p=1.2×10^−6^) in smoking status, resulting in a combined R² of 8.04%. **Fig. 1a** (light gray) shows the additional contribution of parental smoking to models at all thresholds. As thresholds included more SNPs, the contribution of parental smoking decreased and PRS increased; post-hoc tests to examine the relationship in more detail (linear regression) showed that parental smoking was significantly associated with higher PRS at all thresholds (**Fig. 1b**).

#### FTND

PRS for cigarettes per day (Pt=0.2, R =3.15% p=1.82×10−8, **Fig. 1c**) and PRS for smoking cessation (Pt=0.01, R2 =2.01% p=7.18×10−6, **Fig. 1d**) were significantly associated with FTND scores in smokers. Quantile plots depict change in phenotype over PRS in quantiles (**Fig. S2b**,**c**).

## Discussion

The present study demonstrates the informativeness of PRS and highlights their potential utility in the prediction of disease risk. We found that PRS for smoking behaviours were significantly associated with smoking status, and also FTND score in smokers; these associations suggest their importance for use in smoking research as well as the potential to use PRS for purposes of prediction and preventative measures, for example in individuals in the top PRS decile. History of parental smoking was significantly associated with PRS for smoking initiation; overlap with PRS was observed to vary over different PRS score P-value thresholds. Research has found that PRS are able to predict a reliable but modest amount of complex genetic phenotypes and the amount of variance in smoking initiation behaviour phenotypes explained here is in the range of PRS to predict other disease related phenotypes^15^.

Combining SI-PRS with history of parental smoking enabled a better prediction of smoking status. Moreover, the pattern of explained variance over different thresholds, combined with a post-hoc association test, suggests both independent and overlapping components of contribution of parental smoking and PRS. It is of interest to note that in terms of SI, while FH alone explained 3%, the combined model explained up 8%. This highlights the strength of using such a molecular genetic approach, which is more direct than formal genetics. It is also important to note that as the amount of variance explained by the PRS increased, the amount explained by parental smoking decreased. This may suggest that PRS can cover a substantial share of the contributing genetic factors contained in phenotypic parameters such as parental smoking (i.e., family history), which are overall measures that capture not only genetic contributions but also environmental factors; when looking at ‘purely’ biological mechanisms, it may be advantageous to prioritize the use of techniques such as PRS. Used together, these approaches will allow much more detailed analyses and enable exploration of environment-familial loading as well as dissection of biological pathways involved.

We observed that PRS for cigarettes per day and also smoking cessation were associated with FTND score, which points to the ability to identify those who might be at the highest risk of heavy smoking as well and at the same time those who may be able to cease smoking (and are potentially the most responsive to treatment). These results are in line with those of a previous study which used GSCAN summary statistics to generate PRS for amount of smoking and smoking cessation in another European cohort^16^. That study found that in a non-European cohort PRS did not predict smoking behaviours; the generalizability of the results of the present study is also limited by the inclusion of only individuals with European ancestry.

Towards the application of PRS in clinical and practical contexts, other research has found that PRS are informative not only about disease risk but also about time of occurrence; e.g., age of onset ^16,17^. Recent work has also demonstrated that polygenic risk scores are significantly associated with trajectories of problematic substance use and may be informative about developmental progression of disease^18^. It is expected that PRS will be more powerful and potentially approach practical utility with refined phenotypes and PRS based on better characterized and larger GWAS. Moreover, it will be important to extend the approach to different study populations (e.g. of other ancestries). While we constructed the PRS based on risk variants with different thresholds, we also note that the use of PRS does not necessarily have to be limited to this broad approach. PRS can also be refined, e.g., by selecting only variants belonging to specific biological pathways to test whether these play a role in susceptibility for disease or treatment, also with the aim to identify specific subpopulations. Well-defined samples such as the present one offer the possibility to identify relevant pathways using the PRS approach.

Including PRS for smoking initiation in the exploration of an individual’s risk appears to be useful to inform about an individual’s risk of becoming a smoker and may have the potential to inform prevention and intervention strategies. Combining this with FH yielded additional predictive ability, suggesting that the two should be used together. Our findings, serving as a proof of principle, demonstrate this association even in a limited sample size, suggesting that there is more to find when both discovery and target sample sizes increase further.

## Data Availability

All data produced in the present study are available upon reasonable request to the authors.

## Data Availability

The summary statistics used in this study are available at https://genome.psych.umn.edu/index.php/GSCAN (accessed May 2022). The genotype data used in this study are not publicly available due to German data privacy and protection regulations. Summary statistics may be available upon reasonable request. PRSice 2.0 is available at https://choishingwan.github.io/PRSice/

